# Modelling the risk of SARS-CoV-2 infection through PPE doffing in a hospital environment

**DOI:** 10.1101/2020.09.20.20197368

**Authors:** Marco-Felipe King, Amanda M Wilson, Mark H. Weir, Martín López-García, Jessica Proctor, Waseem Hiwar, Amirul Khan, Louise A. Fletcher, P. Andrew Sleigh, Ian Clifton, Stephanie J. Dancer, Mark Wilcox, Kelly A. Reynolds, Catherine J. Noakes

**Affiliations:** School of Civil Engineering, University of Leeds, Woodhouse Lane, Leeds, LS2 9JT, UK; Department of Community, Environment, and Policy, Mel and Enid Zuckerman College of Public Health, University of Arizona, Tucson, AZ, USA; Division of Environmental Health Sciences, The Ohio State University, Columbus, OH, United States of America; School of Mathematics, University of Leeds, Woodhouse Lane, Leeds, LS2 9JT, UK; Department of Respiratory Medicine, St. James’s Hospital, University of Leeds, Leeds, UK; School of Applied Sciences, Edinburgh Napier University, Edinburgh, UK; Department of Microbiology, Hairmyres Hospital, NHS Lanarkshire, G758RG, UK; Healthcare Associated Infections Research Group, Leeds Teaching Hospitals NHS Trust and University of Leeds, Leeds, UK

**Keywords:** SARS CoV-2, COVID-19, PPE, surface contact transmission, quantitative microbial risk assessment (QMRA), hospital infection model

## Abstract

Self-contamination during doffing of personal protective equipment (PPE) is a concern for healthcare workers (HCW) following SARS-CoV-2 positive patient care. Staff may subconsciously become contaminated through improper glove removal, so quantifying this risk is critical for safe working procedures. HCW surface contact sequences on a respiratory ward were modelled using a discrete-time Markov chin for: IV-drip care, blood pressure monitoring and doctors’ rounds. Accretion of viral RNA on gloves during care was modelled using a stochastic recurrence relation. The HCW then doffed PPE and contaminated themselves in a fraction of cases based on increasing case load. The risk of infection from this exposure was quantified using a dose-response methodology. A parametric study was conducted to analyse the effect of: 1a) increasing patient numbers on the ward, 1b) the proportion of COVID-19 cases, 2) the length of a shift and 3) the probability of touching contaminated PPE. The driving factors for infection risk were surface contamination and number of surface contacts. HCWs on a 100% COVID-19 ward were less than 2-fold more at risk than on a 50% COVID ward (1.6% vs 1%), whilst on a 5% COVID-19 ward, the risk dropped to 0.1% per shift (sd=0.6%). IV-drip care resulted in higher risk than blood pressure monitoring (1.1% vs 1% p<0.0001), whilst doctors’ rounds produced a 0.6% risk (sd=0.8%). Recommendations include supervised PPE doffing procedures such as the “doffing buddy” scheme, maximising hand hygiene compliance post-doffing and targeted surface cleaning for surfaces away from the patient vicinity.

**Importance:** Infection risk from self-contamination during doffing PPE is an important concern in healthcare settings, especially on a COVID-19 ward. Fatigue during high workload shifts may result in increased frequency of mistakes and hence risk of exposure. Length of staff shift and number of COVID-19 patients on a ward correlate positively with the risk to staff through self-contamination after doffing. Cleaning of far-patient surfaces is equally important as cleaning traditional “high-touch surfaces”, given that there is an additional risk from bioaerosol deposition outside the patient zone(1).

## Introduction

Severe acute respiratory syndrome coronavirus 2 (SARS-CoV-2) is an enveloped virus which has infected in excess of 10 million people to date and caused more than 700,000 deaths worldwide according to Johns Hopkins University’s COVID-19 Dashboard (2). Inanimate objects known as fomites may host pathogens and have the potential to contribute to infection transmission in healthcare environments. This occurs in viral infection spread (3–5) including COVID-19 (6, 7). There appears to be similarity between persistence of SARS-CoV-1 and 2 on surfaces, with viable virus shown to be present for up to 72 hours (8). This allows an opportunity for exposure through hand-to-fomite contacts, especially if surfaces are heavily contaminated. Although personal protective equipment (PPE) such as gloves, gowns, and masks are worn to protect both patient and healthcare worker (HCW) from exposure, self-contamination during PPE doffing processes (9, 10) poses risks to HCW and enables spread from one patient to another during multiple care episodes. SARS-CoV-2 has been detected on healthcare worker PPE (11) and in the environment of rooms where doffing occurs, providing evidence that errors in doffing could facilitate COVID-19 exposure and transmission.

While SARS-CoV-2 has been detected on PPE and patient surfaces, the relationship between viral RNA concentrations and risk of infection is still unknown(12). Quantitative microbial risk assessments (QMRA) involve the use of mathematical models to estimate doses of a pathogen and subsequent infection risk probabilities. Quantifying infection risk for any given dose can be used to guide intervention decision-making and have been used in other public health contexts, such as in setting water quality standards (13). These typically rely on experimental doses of a microorganism inoculated into healthy participants or mice models in a known quantity. Whether they develop the infection can then be recorded(13). QMRA modelling and surface contact models have been used to evaluate multiple transmission pathways. The role of care-specific behaviours in environmental microbial spread (14) includes the effect of glove use in bacterial spread from one surface to another (15) and evaluating risk reductions through hand hygiene or surface disinfection interventions (16–18). While a strength of QMRA is relating environmental monitoring data to health outcomes, a common limitation has been a lack of specific human behaviour data such as hand-to-face or hand-to-surface contact sequences that result in dose exposures (18, 19). The use of the QMRA modelling framework incorporating care type surface contact patterns before potential self-contamination via PPE doffing will offer insight into infection risks per shift, the importance of a doffing buddy and patient room surface cleaning protocols.

The objective of this study is to relate SARS-CoV-2 concentrations on surfaces to predicted risks of exposure and infection for a single healthcare worker over an 8-hour shift and estimate the effects of doffing mistakes and number of care episodes per shift on infection risk per shift.

## Methodology

This approach combines human behaviour and fomite-mediated exposure models for 19 hospital scenarios, for which concentrations of SARS-CoV-2 on hands and infection risk for a single shift are estimated for a registered nurse, an auxiliary nurse and a doctor. A control scenario was defined as a single episode of care with a SARS-CoV-2 positive individual with an 80% probability of self-contamination during doffing. Eighteen other scenarios covered 3 likelihoods of self-contamination: 5%, 50%, and 80%, x 2 case load conditions: 7 patients (low) vs. 14 patients (high) x 3 probabilities of any given patient being COVID-19 positive: low (5%), high (50%), and a 100% COVID-19 positive ward. During low case load conditions, it was assumed that the number of care episodes per shift would be less than for high load conditions. The assumed number of patient care episodes when PPE is worn per shift for low and high case load scenarios were 7 and 14, respectively, based on a respiratory ward in a university teaching hospital in the UK. The low case load estimate was based on communication with a UK NHS consultant, who tracked the number of gowns used by healthcare workers over a week on a mixed COVID-19 8-bed respiratory ward. All model parameters are described and reported in Table 1. Per scenario, three simulations were run where sequences of hand-surface contacts per care episode were care-specific (IV care, observational care, or doctors’ rounds).

**Table 1.** Model parameters and their distributions/point values

| Parameter | Distribution/Point Value | Reference |
| --- | --- | --- |
| Surface contamination ($C_{RNA}$)<br>(RNA/ swabbed surface area) | For infected patient scenarios<br>Surfaces:<br>Triangular (min=$3.3 \times 10^3$, mid=$2.8 \times 10^4$, max=$6.6 \times 10^4$)<br>Patient:<br>Point estimate: $3.3 \times 10^3$ | (1) |
| Area of any given surface ($A_{surface}$)<br>(cm<sup>2</sup>) | Triangular (min=5, max=195, mid=100) | Assumed |
| Fraction of RNA(infective)<br>assumed to be infectious | Uniform (min=0.001, max=0.1) | Assumed |
| Finger-to-surface transfer<br>efficiency ($\beta$) | Normal (mean=0.118, sd=0.088)<br>Left- and right-truncated at 0 and 1, respectively | (5) |
| Surface-to-finger transfer<br>efficiency ($\lambda$) | Normal (mean=0.123, sd=0.068)<br>Left- and right-truncated at 0 and 1, respectively | (5) |
| Finger-to-mouth transfer<br>efficiency ($TE_{H \rightarrow M}$) | Normal (mean=0.339, sd=0.20)<br>Left- and right-truncated at 0 and 1, respectively | (20, 21) |
| Glove doffing self-contamination transfer<br>efficiency | Uniform (min=$3 \times 10^{-7}$, max=0.1) | (9) |
| $T_{99}$ on Hands<br>(hours) | Uniform<br>(min=1, max=6) | (22, 23) |
| $T_{99}$ on surfaces<br>(hours) | Uniform<br>(min=3, max=120) | (22) |
| Hand hygiene efficacy:<br>alcohol gel (log<sub>10</sub> reduction) | Uniform (min=2, max=4) | (24) |
| Hand hygiene efficacy: soap<br>and water (log<sub>10</sub> reduction) | Normal (mean=1.62, sd=0.12)<br>Left-and right-truncated at 0 and 6, respectively | (25) |
| Fraction of grip ($S_m$ and $S_h$) | For in/out events:<br>Uniform (min=0.10, max=0.17)<br>For patient contacts:<br>Uniform (min=0.04, max=0.25)<br>For other surface contacts:<br>Uniform (min=0.008, max=0.25)<br>For hand-to-face contacts:<br>Uniform (min=0.008, max=0.012) | (26) |
| Total hand surface area ($A_h$)<br>(cm<sup>2</sup>) | Uniform (min=445, max=535) | (19, 27) |
| Dose response curve<br>parameter* $\alpha$ | 0.36 ± 0.25<br>0.12, 19.6 | (28); This<br>study |
| Dose response curve<br>parameter* $\beta$ | 5.94 ± 11.4<br>0.27, 802.1 | (28); This<br>study |

\*Dose response curve parameters are to be used in bootstrapped pairs. Mean ± SD and minimum and maximum are provided to offer context as to the magnitude of these parameters. Bootstrapped pairs are available in supplemental data.

### Healthcare Worker Surface Contact Behaviour Sequences

Fifty episodes of mock patient care were recorded overtly using videography in a respiratory ward side room at St James’ Hospital, Leeds. Mock care was undertaken by doctors and nurses with a volunteer from the research team to represent the patient. While these observations were carried out prior to COVID-19, it is assumed that patient care would be similar for any infected patient, including a COVID-19 patient. Ethical approval for the study was given by the NHS Health Research Authority Research Ethics Committee (London-Queen Square Research Ethics Committee), REF: 19/LO/0301. Sequences of surface contacts were recorded for three specific care types: IV drip insertion and subsequent care (IV, n=17) conducted by registered nurses (RN); blood pressure, temperature and oxygen saturation measurement (Observations, n=20) conducted by auxiliary nurses; and doctors’ rounds (Rounds, n=13). Data from care were used to generate representative contact patterns to model possible sequences of surface contacts by HCWs in a single patient room. HCWs were found to touch surfaces in a non-random manner, insofar that moving from one surface category to another has a hsigher probability than a transition elsewhere. By assigning each surface category a numerical value from 1 to 5, where *Equipment* = 1, *Patient* = 2, *Hygiene areas* = 3, *Near-bed surfaces* = 4, and *Far-bed surfaces* = 5, HCW sequential contact of surfaces can be modelled in terms of weighted probabilities(14).

The movement of a HCW between surfaces is modelled using a discrete-time Markov chain approach (14). Using defined weighted probabilities based on observation of patient care, surface contact by HCW can be simulated based on the property that, given the present state, the future and past surfaces touched are independent. This is termed the Markov property (eq 1):

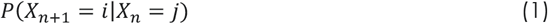

where *X*_*n*_ represents the surface contacted in the *n*^*th*^ event, *i* and *j* are two surfaces, and *P* represents a conditional probability. This is then denoted *P*_*j*→*i*_ for ease of notation. For example, the probability if the HCW is currently touching the table that they will next touch the chair is *P*_*table*→*chair*_ and can be worked out by counting the number of times this happens during care divided by the number of times any surface is touched after the table(29).

Discrete-time Markov chains were fitted to observed care contact sequences using the “markovchainFit” function from the R package *markovchain (version 0*.*7*.*0)*. Separate Markov chains were fitted to IV care, doctors’ rounds and observational care sequences. States included “in” (entrance to the patient room), “out” (exit from the patient room), contact with a far-patient surface, contact with a near-patient surface, contact with a hygiene surface (e.g. tap, sink, soap or alcohol dispenser), and contact with equipment. For each episode of care, the first event was entrance into the patient room. All HCWs wore a gown, gloves, mask and face shield when entering the room. The episode of care ended when an “out” event occurred.

### Exposure Model

Accretion of microorganism on hands from surface contacts has been demonstrated (14) to respond to a recurrence relationship which the concentration on hands after the n^th^ contact, 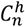, with the concentration on hands,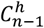, and on the surface involved, 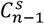, before the contact. See eq. 2.

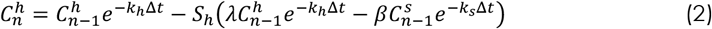

This is an adaptation of the pathogen accretion model (PAM) from King et al. (2015) (14) and a gradient transfer model by Julian et al. (2009) (30). Here, the concentration on hands for contact *n* is equal to the previous concentration on the hand 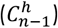 after adjusting for inactivation for the virus on the hand (*k*_*h*_) and surface *k*_*s*_, minus the removal from the hand due to hand-to-surface transfer plus the gain to the hand due to surface-to-hand transfer. Δ*t* is the time taken for an episode of patient care and sampled from a uniform distribution of range 2-20minutes(31). Here, *λ* and *β* represent hand-to-surface and surface-to-hand transfer efficiencies respectively. The fraction of the total hand surface area (*S*_*h*_) is used to estimate how much virus is available for transfer given a concentration of number of viral particles/cm^2^ on the gloved hand and surface.

### Estimating Inactivation on the Hand

Sizun et al. (2000) evaluated the survival of human coronaviruses (HCoV) strains OC43 and 229E on latex glove material after drying. Within six hours, there was a 99% reduction in viral infectivity for HCoV-229E (22). For HCV-OC43, there was a 99% (T_99_) reduction viral infectivity within an hour (22). Harbourt et al. measured SARS CoV-2 inactivation on pig skin with virus remaining viable for up to 8 hours at 37°C (32). We therefore used a uniform distribution with a minimum of 1 hour and a maximum of 8 hours to estimate a distribution of *k*_*h*_ inactivation rates.

### Estimating Inactivation on Surfaces

The decay of the virus causing COVID-19 has been shown to vary under both humidity and temperature but in contrast with previous findings(8), it appears that surface material may not have a significant impact on decay rate(33). We therefore take a conservative approach and use an averaged half-life *τ* estimate for stainless steel and plastic-coated surfaces at 21-23°C(8) at 40% relative humidity; which are 5.63h (95%CI=4.59-6.86h), and 6.81h (95%CI=5.62-8.17h), respectively. We assume a first order decay (eq 3) to estimate inactivation constant *k* which we use here for brevity instead of *k*_*s*_ and *k*_*h*_ in equation 2.

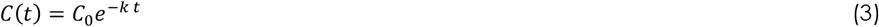

Surface viral concentration C at any given time t then depends uniquely on initial concentration*C*_0_. Where the half-life *τ*, is related to k by: *k*_*s*_ = log(2) / *τ*. Since hospital rooms are made up of a combination of stainless steel and plastic surfaces, we have taken the widest confidence interval as bounds when sampling from a uniform distribution for inactivation rate *k*_*s*_. Inactivation on gloves is assumed to be minimal for the time-scale of a care episode (2-20minutes)(31).

### Fractional Surface Area

For contacts with the door handle during “in” or “out” behaviors, a fractional surface area was randomly sampled from a uniform distribution with a minimum of 0.10 and a maximum of 0.17 for open hand grip hand-to-object contacts (26). For contacts with the patient, a fractional surface area was randomly sampled from a uniform distribution with a minimum of 0.04 and a maximum of 0.25, for front partial finger or full front palm with finger contact configurations (26). For contacts with other surfaces, fractional surface areas were randomly sampled from a uniform distribution with a minimum of 0.008 and a maximum of 0.25, spanning multiple contact and grip types from a single fingertip up to a full palm contact (26).

### Transfer Efficiencies

For contacts with surfaces other than the patient, a truncated normal distribution with a mean of 0.123 and a standard deviation of 0.068 with maximum 1 and minimum 0 was randomly sampled for surface-to-finger(*λ*) transfer efficiencies based on aggregated averages of influenza, rhinovirus and norovirus(5). For patient contacts, transfer efficiencies were randomly sampled from a normal distribution with a mean of 0.056 and a standard deviation of 0.032, left-and right-truncated at 0 and 1, respectively. The mean and standard deviation were informed by transfer efficiencies for rhinovirus measured for direct skin to skin contact (34). Transfer efficiencies from fingers to surfaces (*β*)are assumed to be normally distributed with mean 0.118 and standard deviation 0.088(5).

### Surface Concentrations

If the patient was assumed to be infected, surface contamination levels (RNA/ swab surface area) were sampled from a triangular distribution where the minimum and maximum were informed by minimum and maximum contamination levels reported for surfaces in an intensive care unit ward (1). The median of these was used to inform the midpoint of the triangular distribution (1). For patient contacts, the concentration of virus detected on a patient mask was used as a point value (3.3 x 10^3^RNA/swab surface area) (1). When a patient was not infected, it was assumed contacts with surfaces and with the patient would not contribute to additional accretion of virus on gloved hands.

Surface areas to relate RNA/swabbed surface area to RNA/cm^2^ were not provided by Guo et al. (2020). While a typical sampling size is 100 cm^2^, it may be as small as 10-25 cm^2^ (35–38) and in real-world scenarios, sampling surface areas may be larger or smaller than these depending upon available surface area, ease of access and the contamination magnitude expected. Since the surface areas of these surfaces were not provided, a triangular distribution (min=5, max=195, mid=100) describing the surface area (cm^2^) of surfaces sampled was used to estimate RNA/cm^2^. Not all detected RNA was assumed to represent infectious viral particles. This is a conservative risk approach when utilizing molecular concentration data in QMRA (39). Therefore, concentrations on surfaces *C*^*S*^ (viable viral particles/cm^2^) were estimated by eq 4,

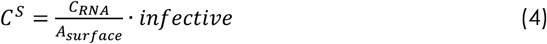

where *C*_*RNA*_ is the RNA/swabbed surface area, *A*_*surface*_ is the surface area (cm^2^) of the surface, and *infective* is the fraction of RNA that relates to infective viral particles (uniform(min=0.001, max=0.1)).

### Estimating Infection Risk

For all scenarios, it was assumed the starting concentration on gloved hands for the first episode of care was equal to 0 viral particles/cm^2^. If gloves were doffed and a new pair was donned in between care episodes, it was assumed the next episode of care began with a concentration of 0 viral particles/cm^2^ on the gloved hands. After each care episode, a number was randomly sampled from a uniform distribution with a minimum of 0 and a maximum of 1. If this value was less than or equal to the set probability of self-contamination during doffing, self-contamination occurred, where the fraction of total virus transferred from the outer glove surface to the hands was assumed to be uniformly distributed between 3 x 10^−5^ % and 10% (9). There was then a 50/50 chance that either hands were washed or sanitized using alcohol gel. If they washed their hands, a log_10_ reduction was randomly sampled from a normal distribution with a mean of 1.62 and a standard deviation of 0.12, (min=0 and max=6) (25). While these are not coronavirus-specific hand washing efficacies they allow for a conservative estimate. If hand sanitizer was used, a log_10_ reduction was randomly sampled from a uniform distribution with a minimum of 2 and a maximum of 4 (24).

To estimate a dose, an expected concentration on the hands after doffing and hand hygiene was estimated, followed by an expected transfer to a facial mucosal membrane during a single hand-to-nose contact after each patient care episode (eq. 5).

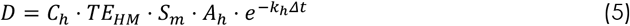

There was a 50/50 chance that either the right or left hand was used for this hand-to-face contact. Here, the transfer efficiency (T_H→M_) of the hand-to-nose contact was randomly sampled from a normal distribution with a mean of 33.90%, and a standard deviation of 20% based on a viral surrogate(21). The fractional surface area of contact (*S*_*m*_) was assumed to equal one fingertip. To estimate this surface area, the minimum and maximum front partial fingertip fractional surface areas were divided by 5 to inform the minimum and maximum values of a uniform distribution (22). The surface area of a hand (*A*_*h*_) was randomly sampled from a uniform distribution with a minimum of 445 cm^2^ and a maximum of 535 cm^2^ (19) and is informed by values from the Environmental Protection Agency, USA’s Exposure Factors Handbook (27). The expected inactivation of virus during this contact assumed a single second contact, and the final *k*_*h*_ value used in the care episode simulation was used. Δ*t* represents the time between doffing and touching the mucosa. 10,000 parameter combinations are obtained for each care type scenario in a Monte Carlo framework.

### Dose-Response

Due to lack of dose-response curve data for SARS-CoV-2, an exact beta-Poisson dose-response curve (40) was fitted to pooled data for SARS-CoV-1 and HCoV 229E, assuming the infectivity of SARS-CoV-2 lies between the infectivity for these two organisms. In eq 6., _1_*F*_1_(*α, α* + *β*, −*d*) is the “Kummer confluent hypergeometric function” and *P(d)* is the probability of infection risk given dose: *d* (eq. 6) (40).

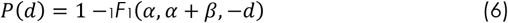

Ten-thousand bootstrapped pairs of *α* and *β* were produced based on a maximum likelihood estimation fit. For each estimated dose, an *α* and *β* pair were randomly sampled, and an infection risk was estimated with eq. 6. The infectious dose for 50% of infections to occur was between 5 and 100 infectious viral particles with a mean of 30; the dose-response curve can be seen in Figure 1.

**Figure 1.**
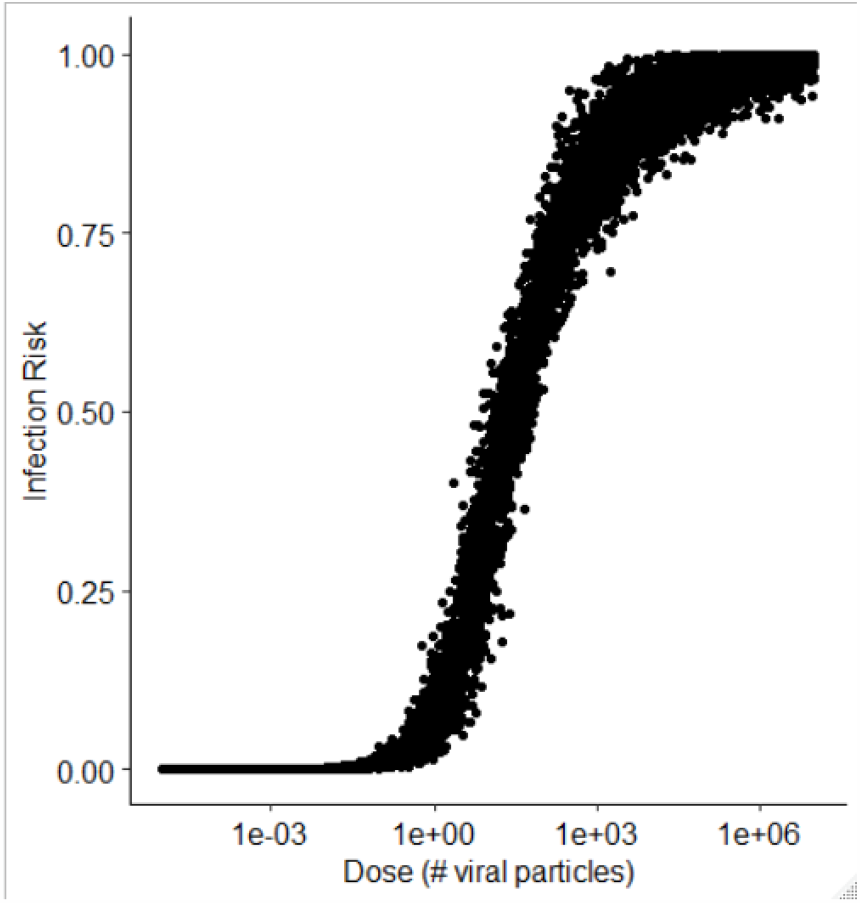
Dose-response risk curve for averaged SARS CoV-1 and Coronavirus 229E response.

### Sensitivity Analysis

Spearman correlation coefficients were used to quantify monotonic relationships between input variables and infection risk. This method has been used in other QMRA studies to evaluate the relationship between model inputs and outputs (30, 41, 42). As not all relationships are monotonic, scatter plots of input variables and associated infection risks were also investigated.

## Results

Surface contact pattern predictions varied by care type. IV care resulted in the highest number of surface contacts (mean=23, sd=10) per episode, whilst observational care and doctors’ rounds had on average 14 (sd=7) and 20 (sd=6) contacts, respectively. A stair plot showing an example HCW surface contact pattern derived from the Markov chain prediction can be seen in Figure 2.

**Figure 2.**
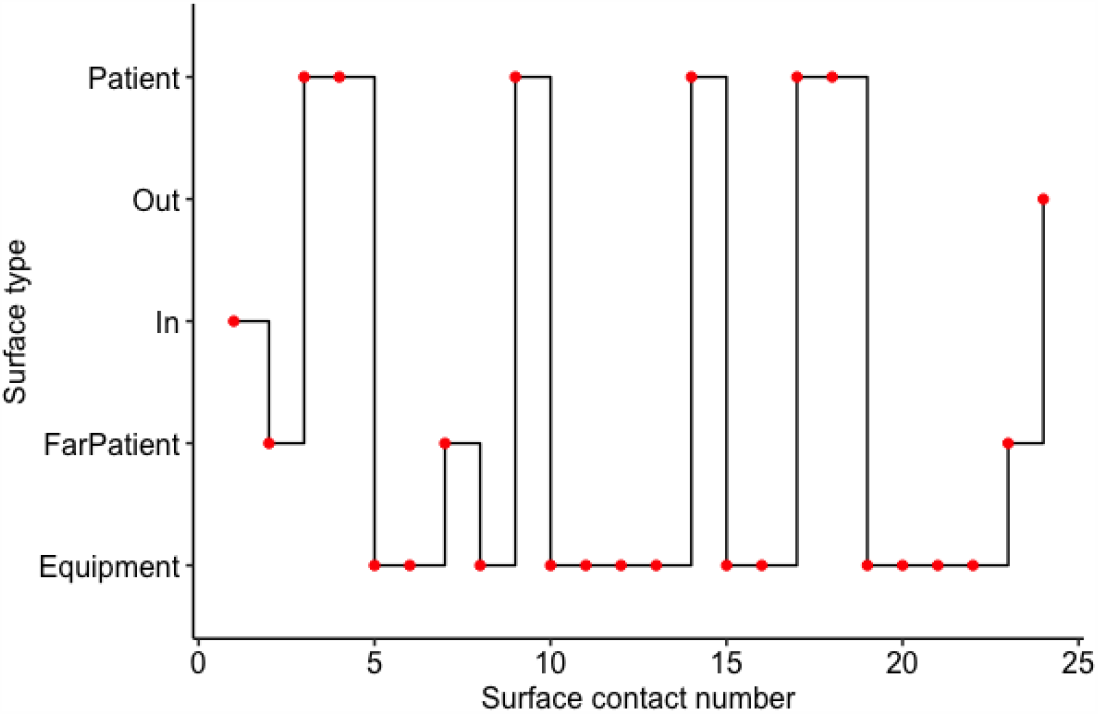
Stair plot of example HCW surface contacts during care.

### Estimated Infection Risks

After a care episode of any type, the risk of becoming infected from a single facial contact ranged from 0 to 29% with a mean of 0.18%. On average, IV care provided the highest risk (1% per shift, p<0.001) due to the number of surface contacts (IV-drip care: 23±10, Doctors’ rounds: 14±7 and Observational care: 20±6). At a 7-patient load, regardless of COVID-19 prevalence, risk was 0.6% whilst doubling patient capacity more than doubled the risk to 1.3% per shift. Figure 3 shows a bar chart with standard deviations for care type, COVID-19 prevalence on the ward and chance of self-contamination following a mistake during doffing.

**Figure 3:**
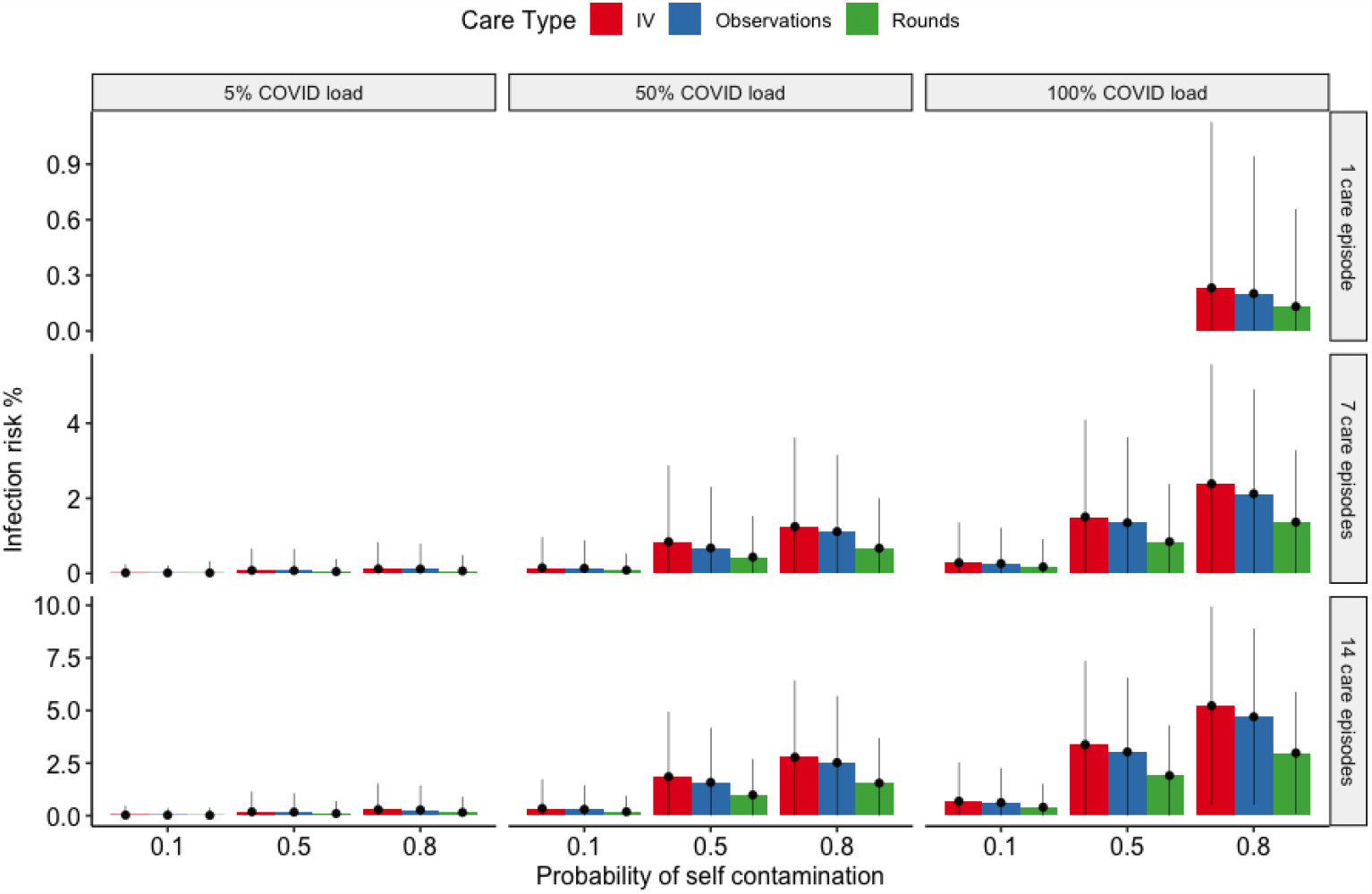
Bar chart showing infection risk % per shift for IV, Observations and doctors’ rounds for different COVID patient loads. Errorbars represent standard deviation of the mean.

Risks relating to COVID prevalence on the ward does not appear to track linearly. HCWs on a 100% COVID ward were less than a 2x more at risk than on a 50% COVID ward (1.6% vs 1%), whilst on a 5% COVID, the risk dropped to 0.1% per shift on average (sd=0.6%).

In terms of most important factor determining risk, Figure 4 shows two dimensional heatmaps of input parameters plotted against predicted infection risk to elucidate correlations. The stronger the correlation, the more influence that parameter has on the output. Surface cleanliness was found to be the single most important factor in determining future risk, with hand-to-mouth/eyes/nose transfer efficiency only half as important (correlation coefficient *ρ* = 0.29 vs *ρ* = 0.12, respectively) (see Table 2). Surface concentration relates to cleaning frequency and hence the control case was run for half the surface bioburden. At double the cleaning frequency, the risk is halved.

**Table 2.** Spearman correlation coefficients of input parameters with infection risk

| Parameter | Spearman Correlation Coefficient |
| --- | --- |
| Concentration on surfaces<br>(viral particles/cm<sup>2</sup>) | 0.29 |
| Transfer efficiency to<br>mouth, eyes, or nose** | 0.12 |
| Transfer efficiency<br>surface to hand | 0.08 |
| Transfer efficiency<br>Hand to surface | 0.06 |
| Inactivation constant for surfaces | 0.05 |
| Fraction of total hand surface area<br>in contact | -0.03 |
| Fraction of RNA relating to infectious<br>particles* | 0.03 |
| Fraction of total hand surface area<br>used in hand-to-face contact** | 0.03 |
| Total hand surface area** | 0.02 |
| Duration (seconds) | 0.02 |
| Inactivation constant for hands | 0.01 |
| Alpha dose response curve<br>parameter | -0.01 |
| Beta dose-response curve<br>parameter | -0.002 |

\*The spearman correlation coefficient represents instances where contacts with surfaces that had non-zero concentrations were made
\*\*The spearman correlation coefficient represents instances in which these parameters were used in a simulation where a contaminated hand-to-face contact was made after doffing

**Figure 4.**
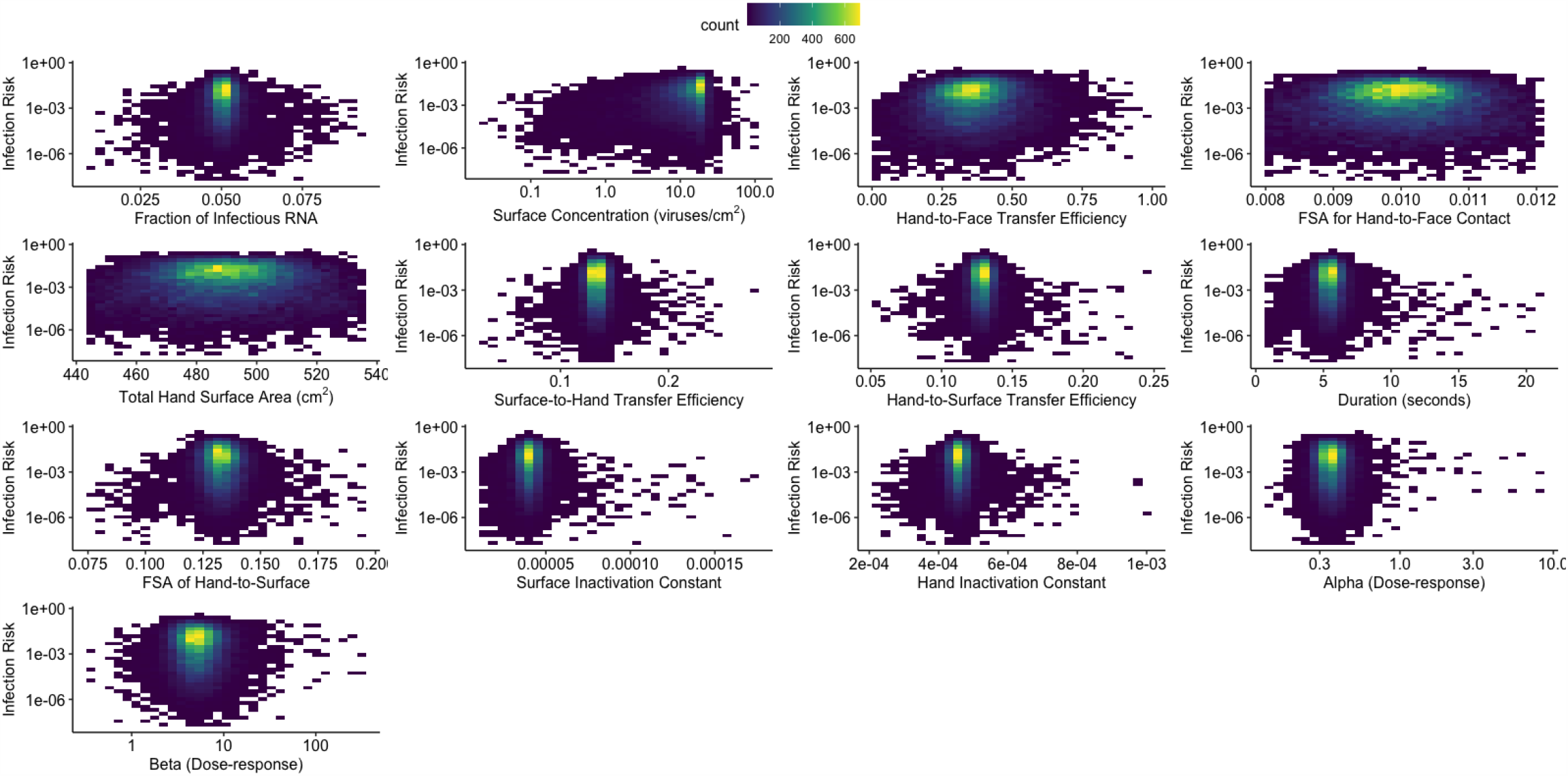
Heatmaps of input parameters plotted against estimated infection risks for scenarios in which doses were greater than zero. FSA= fractional surface area. Count represents the number of simulations resulting in a specified infection risk.

## Discussion

### Key Findings and Generalizability

The model developed in this study indicates that risk of infection from mistakes after doffing PPE is likely to be less than1% for a single shift, even for nurses on 100% patient COVID-19 positive wards. Infection risks vary by care type as greater frequencies of surface contacts directly impact on viral loading on gloves and subsequent self-contamination exposures. The risk increases further if error rates in doffing are high and a high proportion of patients are COVID-19 positive (Figure 1), This highlights the importance of optimal hand hygiene, especially after PPE doffing.

Surface cleanliness was the most important factor in predicting risk regardless of doffing mistake likelihood, highlighting the relevance of frequency of cleaning regimes for managing risk. Halving the surface viral concentration decreased the infection risk 2-fold. Studies have shown that microorganisms can be readily transferred between touch sites in a healthcare environment by routine activities(43). Dispersion of respiratory droplets and aerosols may contaminate less frequently touched surfaces as well, particularly where the patient is undergoing treatment that generates aerosols such as continuous positive airway (CPAP) ventilation. Sampling in COVID wards suggests aerosol deposition is a contributor to surface contamination, as one study has reported deposition at a distance of 3m from the patient(12). Previous experimental work aerosolising a bacteria in an air-conditioned hospital room test-chamber showed that surfaces well outside the patient zone can become contaminated with infectious material (44, 45). Since the observational study underlying the Markov chains reveals that at least 10% of staff contacts impact on such surfaces (excluding door handles), then current lists of high-touch surfaces(46) that had historically been prioritised for cleaning, may need to be revised.

Regardless of the number of COVID-19 positive patients on a ward, notable decreases in predicted infection risk were associated with less self-contamination during doffing. For example, for scenarios involving all COVID-19 patients, the mean infection risk for 10% probability of self-contamination while doffing was 0.4%, while the mean infection risk for an 80% probability of self-contamination while doffing was more than a 420% increase at 2.1% (Table 1). This emphasizes the importance of adequate training for PPE use. Less risk of self-contamination will decrease transmission risks, potentially through a doffing buddy. PPE can be an effective strategy for mitigating exposure if proper doffing techniques are used. In addition to training, improvements in PPE design that enhance safety and expediency of doffing may lower self-contamination rates and therefore improve PPE as a mitigation strategy (47). For example, fasteners or ties on gowns/masks were identified as “doffing barriers,” because it was unclear whether these were to be untied and there were difficulties in reaching these ties. Self-contamination due to gowns and masks were not specifically addressed in this model. It is possible that self-contamination during doffing of items other than gloves could increase potential risks due to incorrect doffing. Shortages of PPE have changed normal practice where PPE is worn on a sessional basis rather than renewed for each patient. This means less doffing and potentially less auto-contamination but may increase the risk of virus transfer within the unit.

In addition to the importance of safe and proper doffing, the results from this computational study also emphasize the importance of surface decontamination and environmental monitoring strategies. The concentration of virus on surfaces was the most influential parameter on infection risk, which is consistent with other surface transmission risk studies (30). Whilst SARS-CoV-2 RNA has been detected on surfaces, one limitation to a molecular approach is the lack of information regarding infectivity. In a recent study by Zhou et al. (2020), no surface samples demonstrated infectivity. However, it was noted that the concentrations of SARS-CoV-2 on surfaces were below the current detection limits for culture methodologies (38). While there are known relationships between cycle threshold values and probabilities of detecting viable virus in a sample (48, 49), it is necessary to know what fraction of detected genome copies relate to viral particles for QMRAs. More data are needed to better understand how molecular concentrations, even concentrations below detection limits, relate to infectivity and subsequent infection risk.

### Limitations

The model in this study only evaluates a surface transmission route while in reality, risks posed to healthcare workers are due combined exposure pathways: air, droplet, person-to-person, and surface transmission. As the model only evaluates surface transmission, these infection risks are likely to be an underestimate of the total risk incurred by healthcare workers over an entire shift. In a study of healthcare workers in a facility in Wuhan, China, 1.1% (110/9684) were COVID-19 positive (50). According to CDC, of 428,295 healthcare personnel for which data were available, 20% (84,035/428,295) were COVID-19 cases (51). However, it is not known how many shifts were associated with these infection rates. Additionally, we assumed that wards with non-COVID-19 patients did not have SARS-CoV-2 contamination on surfaces, due to lack of data on SARS-CoV-2 surface contamination beyond COVID-19 wards or patient rooms. There is potential for asymptomatically infected healthcare workers to contribute environmental contamination, especially when considering the relatively long shedding durations for asymptomatic infections (52). Infected healthcare workers and environmental contamination could be considered in future extensions of this model.

The fact that the proportions of healthcare workers with COVID-19 discussed above are much larger than the infection risks estimated suggest that other transmission routes could drive additional HCW cases. This would include more transmission through airborne routes, or HCW to HCW transmission by asymptomatic cases outside the COVID-19 care environment (53). However, while there continues to be disagreement over the contribution of each route to overall risk, transmission routes influence each other, making them all significant in healthcare environments. For example, surfaces can become contaminated due to deposition of aerosolized virus. Viruses can later be resuspended from surfaces, contributing to air contamination. Future work should extend current models with a multi-exposure pathway approach. This will advance not only our understanding of SARS-CoV-2 transmission but the transmission of pathogens in built environments as a whole.

Finally, a dose-response curve informed by SARS-CoV-1 and HCoV 229E data was utilized, due to lack of SARS-CoV-2-specific dose-response data. We suggest that this therefore is a conservative estimate. Despite limitations related to dose-response, the conclusions from the estimated doses were consistent with insights from infection risk estimates. Increases in probability of contamination between care episodes related to increases in dose and most notably, for scenarios in which more than 5% of patients had COVID-19 (Figure 3, Figure S1).

## Conclusion

We propose a model for predicting risk of infection to healthcare workers from self-contamination during doffing of personal protective equipment over a single shift. The model estimates the quantity of SARS-CoV-2 virus accretion on gloved hands for three types of non-aerosol-generating procedures: IV-care, observations and doctors’ rounds. Once doffing was in progress, staff self-contaminated a fraction of the times based on patient-load fatigue. Three COVID-19 positive patient scenarios (5%, 50% and 100% COVID-19 patients) were investigated amounting to a total of 30,000 parameter combinations allowing us to conduct a “what-if” parametric study and sensitivity analysis. Surface viral concentration was found to be more than twice as important as any other factor whereby highlighting the importance of time-appropriate cleaning. Transfer efficiency from finger to nose was of secondary importance, although hand hygiene following doffing is highly recommended. A doffing buddy could help reduce mistakes regardless of patient numbers and hence is also recommended. Whilst risk from this type of self-contamination sits at around 1% per healthcare worker shift, this highlights that the procedures, if carried out correctly, are generally safe. It is accepted that other routes of transmission will play a significant role in infection propagation.

## Data Availability

Under a Creative Commons Zero v1.0 Universal license (CC-BY), code can be accessed at: https://github.com/awilson12/surface-contam-model-COVID19

## Conflicts of Interest

None to declare

## Acknowledgements

The authors thank Marc P. Verhougstraete, PhD; Christina Liscynesky, MD for their shared expertise and experience that contributed to the development of the modelled scenarios in this study. This research is funded by the Engineering and Physical Sciences Research Council, UK: Healthcare Environment Control, Optimisation and Infection Risk Assessment (https://HECOIRA.leeds.ac.uk) (EP/P023312/1). M. López-García was funded by the Medical Research Council, UK (MR/N014855/1). J. Proctor was funded by EPSRC Centre for Doctoral Training in Fluid Dynamics at Leeds (EP/L01615X/1). Under a Creative Commons Zero v1.0 Universal license (CC-BY), code can be accessed at: https://github.com/awilson12/surface-contam-model-COVID19

## Notes

### Competing Interest Statement

The authors have declared no competing interest.

### Author Declarations

Ethical approval for the study was given by the NHS Health Research Authority Research Ethics Committee (London - Queen Square Research Ethics Committee), REF: 19/LO/0301.

